# Impaired neutrophil-mediated cell death drives Ewing’s Sarcoma in a two years old child with Down Syndrome

**DOI:** 10.1101/2023.05.30.23289664

**Authors:** Serena Peirone, Elisa Tirtei, Anna Campello, Caterina Parlato, Simonetta Guarrera, Katia Mareschi, Elena Marini, Sebastian Dorin Asaftei, Luca Bertero, Mauro Papotti, Francesca Priante, Sarah Perrone, Matteo Cereda, Franca Fagioli

## Abstract

Ewing’s Sarcoma (EWS) has been reported in seven children with Down Syndrome (DS). To date, a detailed assessment of this solid tumor in DS patients is still missing. Here, we characterized a chemo-resistant mediastinal EWS in a 2-year-old DS child, the youngest ever reported case, by exploiting sequencing approaches. The tumor showed a neuroectodermal development driven by the EWSR1-FLI1 fusion. The inherited myeloperoxidase deficiency of the patient caused failure of neutrophil-mediated cell death and promoted genomic instability. In this context, the tumor underwent nearly genome-wide haploidization resulting in a massive overexpression of pro-inflammatory cytokines. Recruitment of defective neutrophils fostered the fast evolution of this EWS.

## Background

Down syndrome (DS) is the most common chromosomal abnormality in Europe, and it is characterized by trisomy of chromosome 21 (Bull 2020; European Commission 2018). DS patients have an elevated risk to develop hematological malignancies (Lee et al. 2016). Conversely, solid tumors are largely underrepresented in these children compared to the euploid population (Satgé et al. 1998, 2013; Hasle et al. 2016; Osuna-Marco et al. 2021). Amongst solid tumors, bone and soft-tissue sarcomas are one of the few histotypes that have been reported in these patients (Osuna-Marco et al. 2021). An extremely small fraction of sarcomas consists of primary mediastinal lesions, a clinically-aggressive neoplasm with poor patient prognosis (Suster 2020). These heterogeneous groups of tumors include small round blue cell sarcomas such as Ewing’s Sarcoma (EWS), which mainly affects children and young adults (Tirtei et al. 2020). This malignancy is characterized by a recurrent chromosomal translocation that fuse an RNA-binding protein of the FET family with a transcription factor of the ETS family, being EWSR1-FLI1 the most common somatic fusion (Grünewald et al. 2018).

So far, seven cases of EWS in young patients with DS (7-19 years old) have been reported and characterized by cytogenetic analyses (Miller 1969; Casorzo et al. 1989; Bridge et al. 1990; Satgé et al. 2003; Kaul, Lotterman, and Warrier 2019). Three tumors (45%) were driven by translocation 11;22 and underwent massive chromosomal changes (Casorzo et al. 1989; Bridge et al. 1990). In particular, these EWSs accumulated amplifications rather than deletions, with recurrent gains of chromosome 8 and 14 (Casorzo et al. 1989; Bridge et al. 1990). The authors of these studies hypothesized an involvement of the constitutional trisomy 21 in driving the disease, implicating the proto-oncogenes ETS1 and ETS2 as oncogenic drivers (Casorzo et al. 1989; Bridge et al. 1990). However, being based on cytogenetic assays, these studies lack a comprehensive molecular characterization of EWS in DS patients.

Here, we comprehensively characterized a mediastinal EWS in a 2-years-old child with DS. Using whole exome and transcriptome sequencing, we highlighted the complex genomic architecture of the EWS characterized by the clonal EWSR1-FLI1 fusion. We identified an inherited rare mutation causative of myeloperoxidase deficiency leading to impairment of neutrophil-mediated cell death and promoting genomic instability. In this background, the tumor genome underwent nearly haploidization resulting in a pro-inflammatory environment. Recruitment of defective neutrophils fostered the fast evolution of the tumor. Our results elucidate the genetics and the predisposing mechanisms of a solid tumor in a young DS patient with possible impacts on their clinical management.

## Results

### Clinical history

A two years old male child affected by DS was presented with a three week history of dyspnea, inspiratory stridor, and episodes of cyanosis with crying (Figure 1). Transthoracic echocardiographic assessment showed a retrocardiac parenchymal mass and massive pericardial effusion, with initial sign of cardiac tamponade. Urgent ultrasound-guided pericardiocentesis was required even if complicated by a cardiac arrest. Sternotomy was then performed with evidence of tumor capsule rupture and bioptic samples of the tumor mass were collected for pathological examination. After stabilization, total body computed tomography (CT) scan revealed a solid heterogeneous mass (8.5 cm x 8 cm x 6 cm) causing deviation of the trachea and the mediastinal vascular structures, with associated right jugular vein thrombosis (Figure 1, upper left panels).

**Figure 1.**
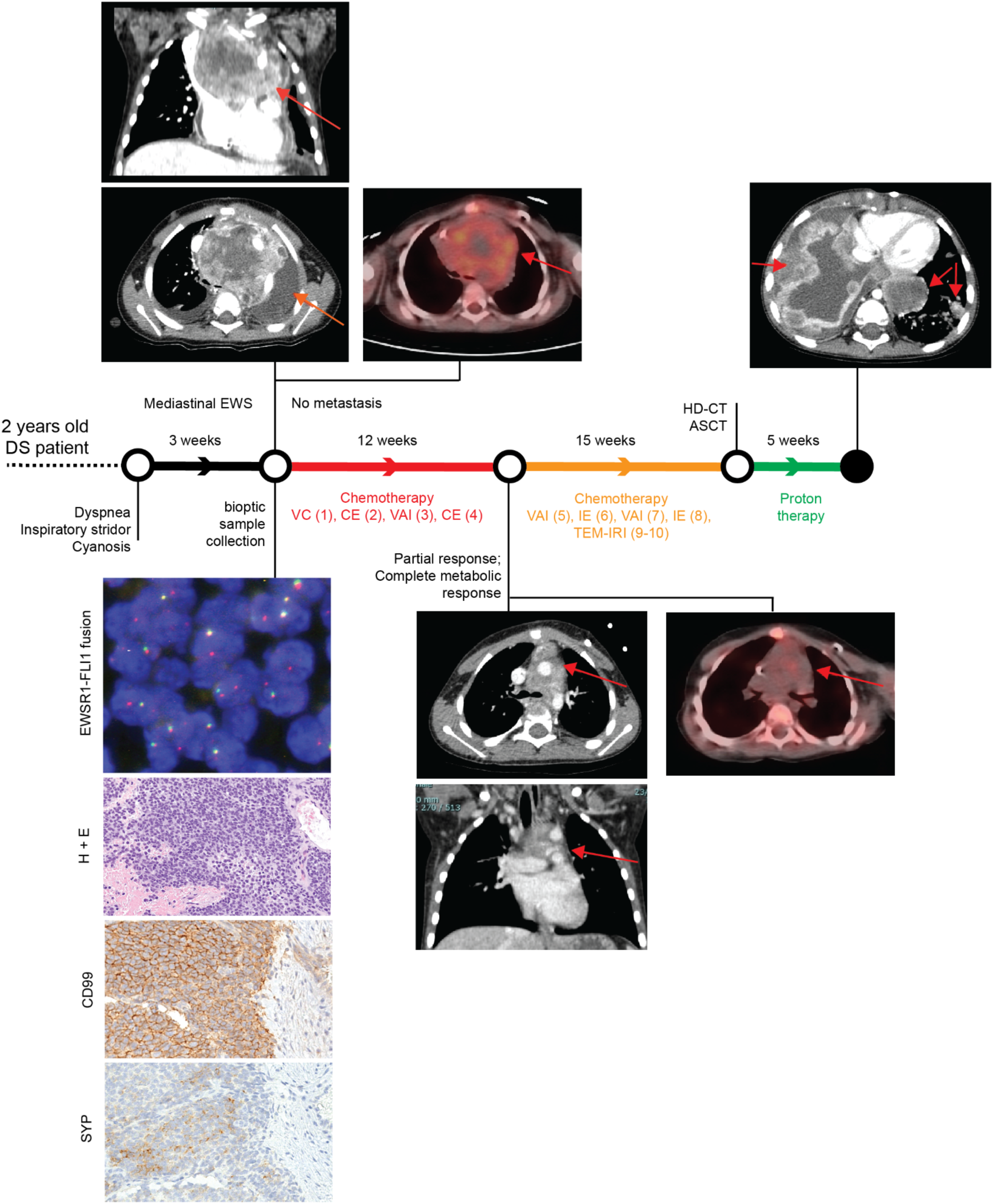
Patient clinical history. Patient history is reported with regard to diagnostic and therapeutic procedures along the time bar. Images from Thoracic CT and PET-CT at diagnosis, after first chemotherapeutic treatment, and after proton therapy (only CT) are shown. Evaluation of EWSR1 translocation t(22q12) by FISH, Hematoxylin and Eosin (H&E), and CD99 immunohistochemical images are reported in the bottom left corner. Magnification 200x. VC= Vincristine (1.4 mg/sqm) + Cyclophosphamide (850 mg/sqm). CE= Cyclophosphamide (4g/sqm) + Etoposide (600mg/sqm). VAI=Vincristine (1.4mg/sqm) + Adriamycin (90mg/sqm) + Ifosfamide (9gr/sqm). IE= Ifosfamide (9gr/sqm) + Etoposide (300mg/sqm). VAC=Vincristine (1.4mg/sqm) + Adriamycin (80mg/sqm)+ Cyclophosphamide(1.2g/sqm). TEM-IRI= Temozolomide (100mg/sqm/day) + Irinotecan (50mg/m2/day). HD-CT/ASCT = High dose chemotherapy and autologous stem cell transplantation (conditioning regimen: Treosulfan (10g/sqm/day x 3 days) + Melphalan (140mg/sqm/day x 2 days).

Histopathological examination detected a small blue round cell tumor (Figure 1, left bottom panels), which was strongly positive for CD99 by immunohistochemistry, thus suggesting an Ewing’s Sarcoma (EWS) (Grünewald et al. 2018). The diagnosis of EWS was supported by the identification of a EWSR1 translocation (22q12.2) using fluorescence *in situ* hybridization (Zöllner et al. 2021). After informed consent signed by the parents, the patient was enrolled into the Italian pediatric sarcoma genomic study SAR-GEN_ITA aiming at profiling its inherited and somatic alterations (ClinicalTrials.gov id: NCT04621201).

A general disease staging was carried out within 72 hours from the histological diagnosis with bilateral bone marrow aspiration and positron emission tomography CT (PET-CT) scan following the European Bone Sarcoma Guidelines (Strauss et al. 2021). The results confirmed the presence of a locally advanced tumor without distant metastasis (Figure 1, upper left panels). A multi-agent induction chemotherapy regimen was delivered to the patient. A first chemotherapeutic cycle of Vincristine and Cyclophosphamide was tailored according to the unstable clinical condition of the patient. Due to cardiac surgical intervention Adriamycin was omitted to avoid adjunctive toxicity. Conversely, Cyclophosphamide was considered more tolerable than Ifosfamide. After the first chemotherapeutic cycle, the patient obtained a clinical benefit with a fully-stabilization of clinical condition without any new dyspnea episode. Therefore, the induction treatment proceeded with three more chemotherapeutic cycles every 21 days: two cycles with Vincristine, Adriamycin and Ifosfamide and one cycle with Carboplatin and Etoposide (Figure 1). A complete radiological tumor response was assessed at the end of the induction period and it evidenced a partial response according to RECIST 1.1 (Schwartz et al. 2016) with a tumor shrinkage of 47% and a complete metabolic response at PET-CT as previously described (Mp et al. 2003; Hicks and Lau 2009) (Figure 1, middle panels). Nevertheless, a complete surgical tumor resection was not feasible. Hence, the patient received additional chemotherapy treatment alternating six poli-chemotherapeutic cycles every 21 days (Figure 1). Next, a consolidation therapy was performed employing a high dose chemotherapy regimen with Treosulfan and Melphalan followed by autologous peripheral stem cell infusion (Figure 1). Again, being the complete surgical excision impracticable, the patient received proton therapy (cumulative dose of 54 Gy in 30 fractions) as local treatment.

Despite persistent evidence of a stable and not metabolically active disease, 16 months after the initial diagnosis, the patient developed disease progression with a massive and rapidly evolving pulmonary involvement that led to patient *exitus* (Figure 1).

### The patient carries a rare damaging germline SNPs in the myeloperoxidase *MPO* **gene**

To determine inherited pathogenic predisposition of the DS patient, we performed whole exome sequencing (WES) on DNA extracted from peripheral blood reaching an average depth of coverage of 66x. We identified germline single nucleotide polymorphisms (SNPs) from sequenced reads and used such information to assess chromosomal anomalies (see Methods). To assess possible inherited changes in chromosome copies, we inspected the variant allele frequency (VAF) distribution of germline SNPs. In particular, shifts of the VAF distribution from the expected peaks of heterozygosity (VAF = 50%) and homozygosity (VAF =100%) are informative of the presence of copy number changes (Cereda et al. 2016). As a result we confirmed the trisomy 21 in this patient (Supplementary Figure 1A).

We next sought to determine additional hereditary conditions that could be associated with, or predispose to, the onset of EWS. To do so, we focused on germline SNPs that are rare in the general population (*i.e.* minor allele frequency <0.001, see Methods), thus most likely to be associated with diseases (Cereda et al. 2016). Out of 6,596 rare germline SNPs, we selected 879 defined as most likely deleterious by the Combined Annotation Dependent Depletion (CADD) algorithm (Kircher et al. 2014) (*i.e.* CADD13 PHREAD score ≥ 10, see Methods). Of these, 17 deleterious SNPs were classified as pathogenic or as variants of uncertain significance (VUS) from at least one of two tools for clinical interpretation of genetic variants, namely ClinVar (Landrum et al. 2020) and Intervar (Q. Li and Wang 2017) (Supplementary Table 1). Amongst these rare deleterious SNPs, the MPO c.2031-2A>C splicing mutation was the only one reported as ‘pathogenic’ by both resources. This rare splicing mutation is known to be causative of myeloperoxidase deficiency (Marchetti et al. 2004). Indeed, by performing conventional splice strength analysis, we predicted a high potential to disrupt the native 3’ splice sites at intron 11 and exon 12 junction (Shamsani et al. 2019) (Supplementary Figure 1B).

### The EWS presented high mutational load and near-haploidization

To assess the somatic alterations that characterize this mediastinal EWS, we extracted genomic DNA from tumor tissue collected at diagnosis and performed WES. We sequenced the exome at an average depth of coverage of 62x and called single base substitutions (SBSs) and small insertions/deletions (ID). We compared variant calling results between tumor and normal samples to identify somatic mutations.

Overall, the SBS landscape of the tumor was characterized by a prevalence of C>T and T>[C/G] substitutions (Figure 2A-B). C>T and T>G substitutions were in the context of G base at the immediate 3’ (*i.e.* N[C>T]G) and of A[T>G]G trinucleotides, respectively. Conversely, T>C substitutions did not present any evident design. The C>T and T>C/G mutational patterns were recapitulated by the known COSMIC SBS1 and SBS5 signatures, respectively (Figure 2B-D and Supplementary Figure 1C). Both mutational signatures have been recurrently found in pediatric cancers (Thatikonda et al. 2023). While SBS5 etiology is of unknown etiology, SBS1 is indicative of deamination of 5-methylcytosine (5mC) to thymine (Thatikonda et al. 2023). The ID signatures presented a more skewed distribution, mainly characterized by single base T insertions and deletions in long thymine homopolymers, as well as small deletions in repeated regions (Figure 2C). This pattern recapitulated a combination of COSMIC ID2, ID12, and ID1 signatures (Figure 2C-D and Supplementary Figure 1D). ID1 and ID2 defined the single base T insertions or deletions at T stretch repeats, whereas ID12 summarized the small deletions at repeated regions (Supplementary Figure 1D). Similarly to SBS1, ID1 and 2 have been recurrently found in pediatric cancers and associated with DNA damage induced by replication slippage (Thatikonda et al. 2023). Although ID12 has been previously identified in pediatric patients with brain tumors (Thatikonda et al. 2023), its etiology is unknown.

**Figure 2.**
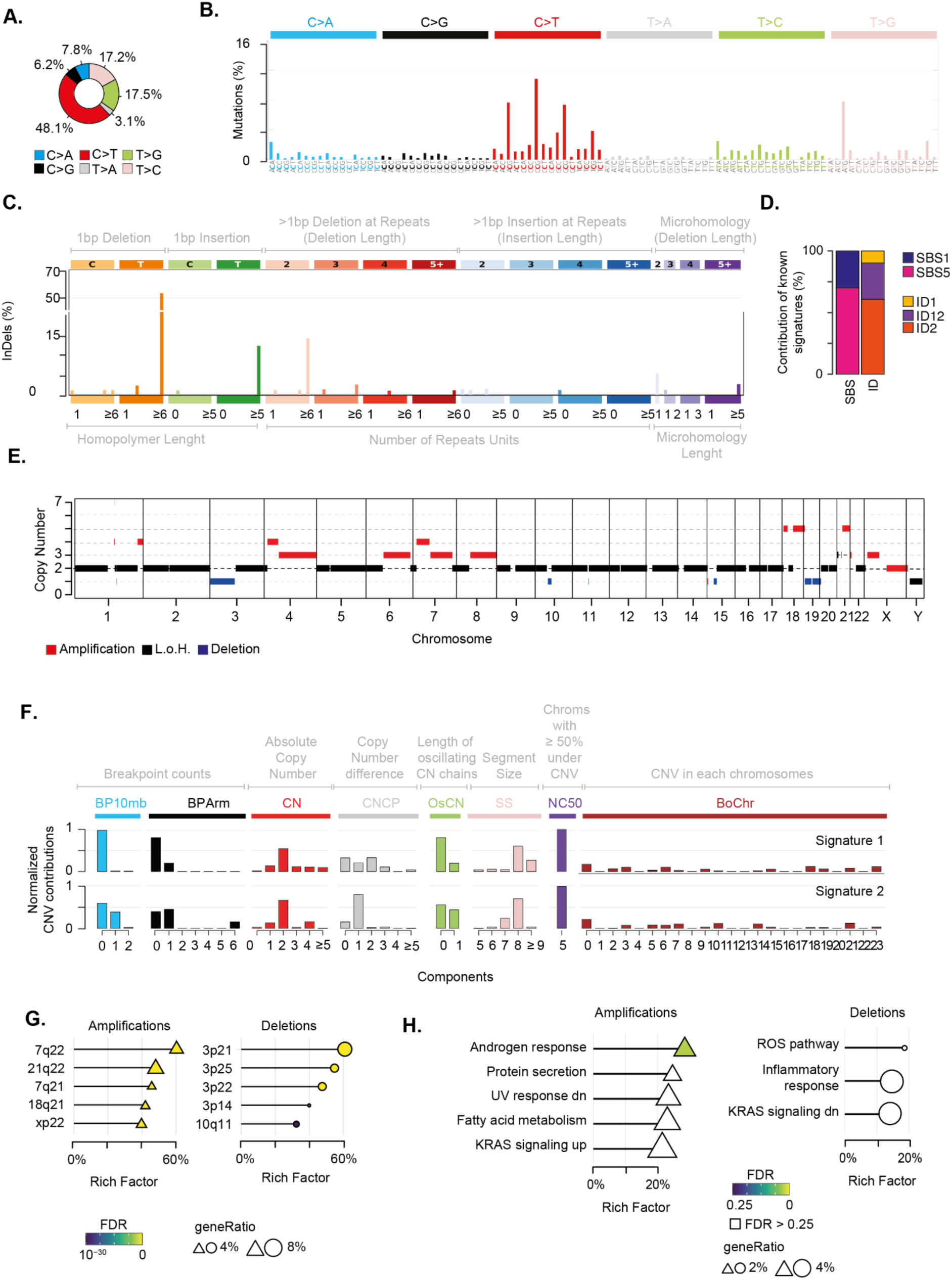
Genomic alterations characterizing the mediastinal EWS. (A) Pie chart depicts the fraction of somatic single base substitutions (SBSs). (B) Most representative mutational SBS signature. (C) Most representative mutational ID signature. (D) Barplot shows the contribution of COSMIC SBS and ID signatures to the most representative signatures detected in the EWS. (E) Chromosomal regions undergoing somatic copy number alterations. (F) Most representative mutational CNV signature. BP10MB = breakpoint count per 10 Mb. BPArm = breakpoint count per chromosome arm. CN=copy number of the segments. CNCP = difference in copy number between adjacent segments. OsCN = lengths of oscillating copy number segment chains. SS = log10 based copy number segment size. NC50 = minimal number of chromosomes with 50% copy number variation. BoChr = burden of chromosome. (G-H) Over representation analysis performed on genes undergoing CNVs relative to chromosomal bands (G) and Hallmark gene sets (H). Shape size indicates the fraction of CNV genes in each pathway (i.e. geneRatio). The Rich Factor represents the fraction of genes in each pathway undergoing CNVs. Color key represents the statistical significance (FDR) of the enrichment. Only top-5 enriched pathways (FDR<0.1), if any, are shown and sorted by statistical significance.

We then inspected the mutational landscape to identify possible driver alterations. In particular, we selected “nonsilent” alterations that were likely to impair the function of the encoded protein (see Methods). These somatic variants accounted for a tumor mutational burden of 2.15, which was in the range of highly mutated pediatric tumors (Gröbner et al. 2018). Out of 142 nonsilent mutations, we identified eight putative driver alterations (Supplementary Table 2). Four of them were marked as “highly deleterious” by the CADD algorithm (*i.e.* CADD13 PHREAD score ≥ 20) affecting known cancer driver genes. In particular, NOTCH2 H107P and BCR T1127S variants were almost clonal (*i.e.* present in all somatic cells), whereas EPHA7 L564F and MTOR S920F were subclonal alterations being present in around 35% of cancer cells. CancerVar classified these mutations as VUS (Q. Li et al. 2022). Nonetheless, NOTCH2 H107P and EPHA7 L564F predicted by CancerVar as “oncogenic” variants with the highest accuracy (*i.e.* Oncogenic Prioritization by Artificial Intelligence (OPAI) score > 0.84).

Next we assessed the chromosomal status of the tumor. By profiling copy number variations (CNVs) on tumor and normal samples we identified regions undergoing somatic alterations (see Methods). We found that 29% of the genome had undergone chromosomal changes, with the majority (22%) being amplifications (Figure 2E and Supplementary Figure 1E and Supplementary Table 3). Furthermore, our analysis revealed that the tumor had a ploidy of 1.4. Therefore, in absence of consistent genomic losses, these findings suggest that the tumor underwent genome-wide massive loss of heterozygosity (LOH) driven by near haploidization.

The analysis of copy number signatures revealed two closely related patterns characterized by (*i*) few arm and focal level breakpoints, (*ii*) a low absolute copy state with small differences between adjacent segments, and (*iii*) large alterations of approximately 100 mega base pairs hitting five chromosomes for more than 50% of their length (Figure 2F). To assess whether CNV localized on specific chromosome regions, we measured the over-representation of genes undergoing CNVs on 278 chromosomal bands (Subramanian et al. 2005). We found that 21q22 and 3p21 regions were the most enriched bands for amplified and deleted genes, respectively (Figure 2G and Supplementary Table 4). In 21q22 we detected amplification of five cancer genes, including the transcription factors ERG and RUNX1 and the RNA binding protein U2AF1. These three genes have been reported as driver genes in pediatric cancers (Ma et al. 2018). It is worth noting that amplification of 21q22 reveals the gain of one copy of the transcription factor ETS2, which has been previously suggested to play a tumorigenic role in these patients (Bridge et al. 1990; Hasle 2001). To identify the biological processes affected by chromosomal changes, we evaluated the over-representation of genes undergoing CNV in a list of 50 Hallmark gene sets. This list defines specific biological states displaying coherent expression (Liberzon et al. 2015; Subramanian et al. 2005). Although not reaching stringent cutoff for multiple test correction, amplifications preferentially affected genes in the androgen response, UV response, protein secretion and metabolism of fatty acids pathways (Figure 2H and Supplementary Table 5). Conversely, deletions impaired preferentially genes in immune-related and reactive oxygen species (ROS) pathways. Interestingly, we found an enrichment for amplifications and deletions in genes that are known to be regulated by the activation of the proto-oncogene KRAS.

Finally, to select putative drivers undergoing CNVs with the greatest accuracy, we exploited the expectation-maximization probability that a gene belongs to a specific copy number state provided by EXCAVATOR2 (D’Aurizio et al. 2016). We identified ten genes with a probability greater than 0.9 to undergo a specific alteration (Supplementary Table 6). Amongst these candidate genes, we found a one-copy amplification of the proto-oncogene MET.

### The EWS derives from neuroectoderm differentiation

Given the complex genomic landscape, we sought to investigate the transcriptomic profile of the EWS. To do so, we extracted total RNA from tumor tissue collected at diagnosis and performed deep RNA sequencing (∼54 Million reads). Firstly, we mapped all gene fusions and identified the EWSR1-FLI1 fusion resulting from translocation t(11;22) as the major oncogenic event (Figure 3A and Supplementary Table 7). The expression of the EWSR1-FLI1 protein induces expression of neuroectodermal differentiation markers (Lin, Wang, and Lozano 2011). In this light, we collected five gene signatures of embryogenesis states (*i.e.* ectoderm, endoderm, mesoderm, neuroectoderm, neuromesoderm) (Messmer et al. 2019; Grosswendt et al. 2020) and measured the cumulative expression of genes in these lists. The neuroectoderm signature was the most expressed compared to the others, thus corroborating the neuroectodermal origin of the EWS driven by the EWSR1-FLI1 fusion (Figure 3B).

**Figure 3.**
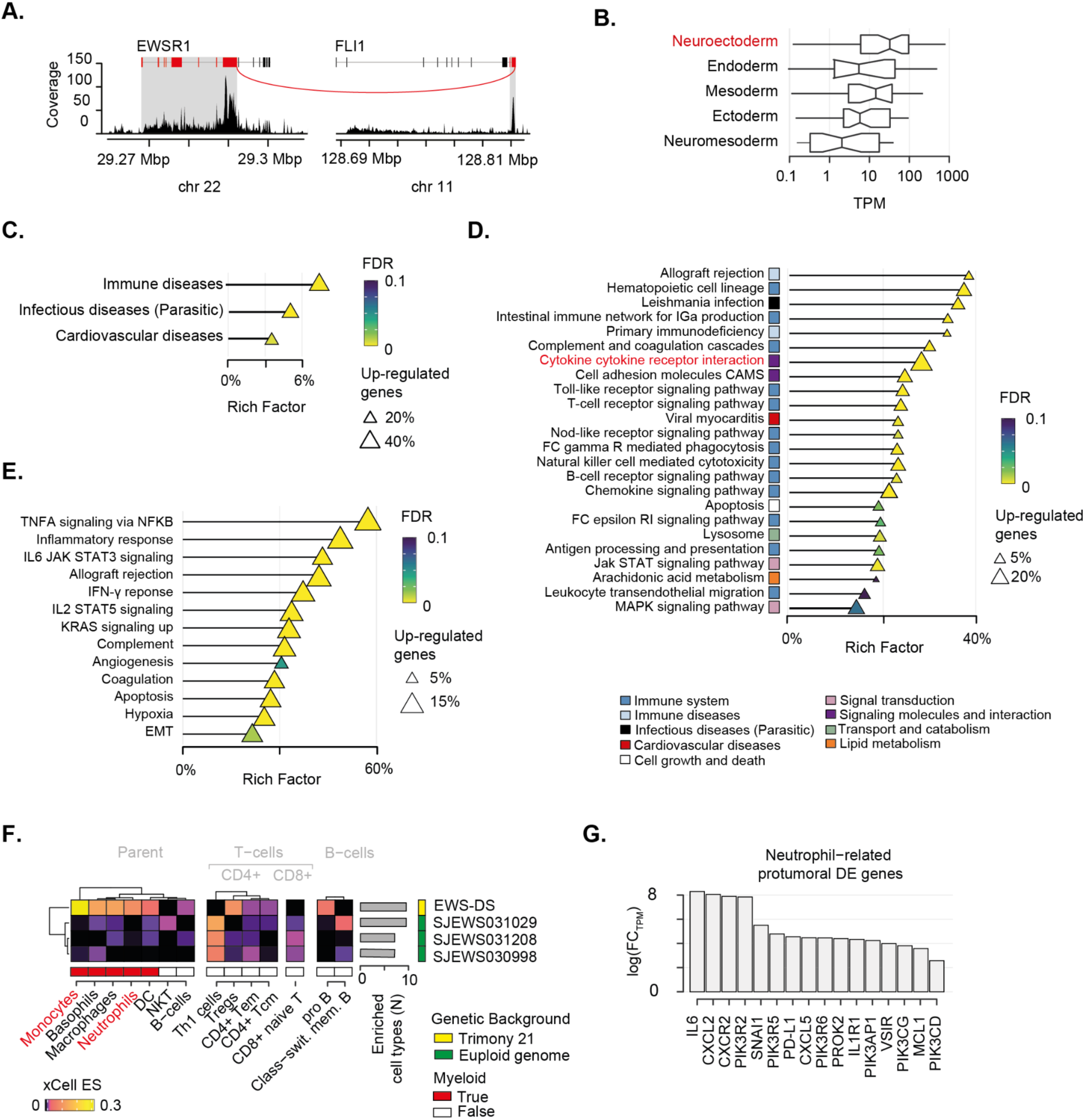
Transcriptomic landscape of the mediastinal EWS. (A) EWSR1-FLI1 fusion breakpoint detected by RNA-seq. Distribution of sequenced reads (i.e. coverage) is shown. Red line indicates the breakpoint of the fusion. (B) Boxplot depicts the cumulative normalized expression levels of genes defining embryogenesis states. (C-E) Over representation analysis performed on differentially expressed (DE) genes relative to KEGG superfamily of gene sets (C), KEGG individual gene set (D), and Hallmark gene set (E). Shape size indicates the fraction of DE genes in each pathway. The Rich Factor represents the fraction of genes in a pathway that are differentially expressed. Color key represents the statistical significance (FDR) of the enrichment. Only enriched pathways (FDR<0.1), if any, are shown and sorted by statistical significance. No enrichment found for down-regulated genes. (F) Heatmap shows immune-cell-specific xCell enrichment scores for the mediastinal EWS and EWSs from euploid patients. Right annotation heatmap depicts the number of enriched cell types for all tumors. (G) Barplot shows fold-change in expression levels in logarithmic scale of neutrophil-related pro-tumoral genes found as DE in the mediastinal sarcoma compared to the other EWSs.

### The EWS-DS microenvironment is characterized by over-represented neutrophil recruitment

To gain insights into the transcriptional programmes that characterized this mediastinal EWS, we collected gene expression data of three additional EWSs from euploid patients of 2-3 years old that were available at the St Jude database (see Methods) (McLeod et al. 2021). We specifically selected children with comparable age of the DS patient and used these data as a baseline for the gene expression comparisons. By performing differential gene expression analysis (see Methods), we identified 2,124 upregulated and 103 downregulated genes (Supplementary Table 9) in the EWS of the DS patient (hereafter referred as EWS-DS) compared to the other EWSs of the euploid cohort (Supplementary Figure 2). We then evaluated the over-representation of these differentially expressed genes in a list of 158 gene sets from the Kyoto Encyclopedia of Genes and Genomes (KEGG) (Subramanian et al. 2005) to identify the altered biological processes characterizing the EWS-DS transcriptome. We found that up-regulated genes were significantly involved in immune-and infectious-disease-related pathways (Figure 3C and Supplementary Table 10), thus corroborating the role of inflammation in our patient. Conversely, the small amount of down-regulated genes were significantly implicated in translation processes (Supplementary Figure 2C and Supplementary Table 10). By performing the over-representation analysis at single gene set level, we found that most of the significantly altered pathways had a clear connection with immune response (Figure 3D and Supplementary Table 10). In particular, a large fraction (∼31%) of the cytokine–cytokine receptor interaction pathway was significantly up-regulated. This proportion accounted for more than 20% of the total differentially expressed genes, indicating a crucial pressure towards the activation of inflammatory response. We orthogonally evaluated the over-representation of up-regulated genes in specific biological states using the Hallmark gene sets (Liberzon et al. 2015; Subramanian et al. 2005). Again, we found a clear enrichment of differentially expressed genes in immune related pathways (Figure 3E and Supplementary Table 10). Specifically, the majority (59%) of the tumor necrosis factor alpha (TNFA) signaling cascade activated by the NF-κB pathway was up-regulated. Similarly, a large fraction of other immune-related protumorigenic pathways, such as IL6-Jak-STAT3, IL2-STAT5, and Interferon gamma (IFN-*γ*) signaling cascade, was overexpressed.

We sought to assess how this inflammatory signature reflected on the EWS-DS immune microenvironment. To do so, we deconvoluted gene expression profiles of the four tumors using xCell (Aran, Hu, and Butte 2017). This algorithm provides an enrichment score for each cell type in each sample that is comparable across conditions. Out of 35 immune cell types, ten were enriched in the EWS-DS as compared to the euploid controls (Figure 3F). Amongst these, myeloid cells such as monocytes and neutrophils were strongly over-represented in the EWS-DS. In light of this evidence, we assess the expression levels of 34 genes that are known markers of the tumorigenic role of neutrophils (Hedrick and Malanchi 2022) (Supplementary Table 8). Overall, 47% of these neutrophils-related tumorigenic markers were significantly differentially expressed in the EWS-DS compared to the other EWSs (Figure 3D). Interestingly, markers of neutrophil trafficking and recruitment during inflammation, such as *IL6*, *CXLC2*, and *CXCR2* showed the highest fold change of expression (McLoughlin et al. 2003; G. Wang et al. 2021).

## Discussion

In this study, we extensively characterized the genetic and transcriptomic landscape of a mediastinal EWS in a two-year old patient with Down’s Syndrome. We showed that this solid tumor had developed a rare genomic architecture likely in the background of inflammation. This condition originated from inherited predisposition of the patient and promoted by the tumor. Our results revealed the putative defective role of neutrophils in fostering the fast evolution of this solid tumor. Since no specific guidelines exist for the management of solid tumors in DS patients, these findings underline the need for rapid genomic screening to extend our understanding of these rare diseases and, eventually, inform on the most appropriate clinical decisions.

Our genomic screening showed the presence of a rare pathogenic splicing variant in *MPO* (c.2031-2A>C) that is responsible for myeloperoxidase deficiency (MPOD) (Marchetti et al. 2004). MPOD is a primary immunodeficiency characterized by a decreased MPO activity in neutrophils (Marchetti et al. 2004; Klebanoff 2005). These myeloid cells are emerging as regulators of cancer development (Hedrick and Malanchi 2022), especially in case of rare malignancies such as synchronous tumors (Cereda et al. 2016). In physiological conditions activated neutrophils release reactive oxygen species (ROS) and MPO to promote cell death (Hedrick and Malanchi 2022). MPO regulates ROS production by catalyzing the assembly of hydrogen peroxide (H_2_O_2_) with halide ions to produce hypohalous acids (Davies et al. 2008). These agents are important for MPO-mediated innate immune response. Loss of MPO leads to accumulation of H_2_O_2_ that amplifies DNA damage and activation of error-prone non-homologous end-joining repair, thereby promoting tumorigenesis (Kongkiatkamon et al. 2022). Therefore, impairment of neutrophil-mediated cell death driven by MPOD may have favored tumorigenesis in the DS patient via increased genomic instability.

Our analyses on somatic alterations corroborates this scenario. We identified age-related mutational signatures (*i.e* SBS1, ID1, and ID2) that characterize pediatric tumors (Thatikonda et al. 2023). The mutational processes underlying these signatures arise from errors that are not repaired during DNA replication at mitosis (Alexandrov et al. 2020). Specifically, the number of SBS1 substitutions mirrors how many mitoses a cell has undergone (Alexandrov et al. 2015). Similarly, ID1 and ID2 mutational signatures result from defects in the DNA mismatch repair (Alexandrov et al. 2020; Thatikonda et al. 2023). These genomic-instability-related signatures coherently describe the high mutational load of this pediatric sarcoma. Therefore, this hyper-mutability may reflect the elevated DNA damage repair levels induced by MPOD occurring during mitosis (Pedersen R et al. 2016; Kongkiatkamon et al. 2022; Hedrick and Malanchi 2022).

Driven by canonical EWSR1-FLI1 gene fusion, the EWS evidenced massive genomic instability, reaching nearly genome-wide haploidization. This is an extremely rare phenomenon whereby the funder clone likely undergoes extensive chromosome loss during mitosis leading to a nearly haploid genome. Near-haploidization has been reported in rhabdomyosarcoma and leiomyosarcomas, and associated with a prominent inflammatory component (Arbajian et al. 2018; Walther et al. 2016). Again, the oxidative DNA damage driven by MPOD may have contributed to the catastrophic near-haploidization of the EWS. Furthermore, somatic chromosomal losses impaired preferentially genes in immune-related and ROS pathways. Therefore, this finding suggests an additional impairment of inflammatory response among the surviving clones.

Genome instability is a known feature of DS patients and there is an open debate on its contribution to cancer progression (Nižetić and Groet 2012). We found that regions of chromosome 21 and 3 (*i.e.* 21q22 and 3p21) were hotspots of amplified and deleted genes, respectively. The possible role for constitutional trisomy 21 in EWS development in DS patients has been hypothesized relying on the presence of oncogenes such as ETS2 on 21q22 (Bridge et al. 1990; Hasle 2001). Here we found that the acquisition of one copy of the *ETS1* locus led to a significant increase of *ETS1* expression in the EWS-DS compared to other EWSs from euploid patients (FC=2.58; FDR=0.037). Furthermore, we identified the amplification of the proto-oncogene MET, a recurrent driver of resistance in multiple solid tumors (Wood et al. 2021). It has been recently shown that MET induced by tumour-derived tumour necrosis factor (TNF)-α promotes anti-tumorigenic activities in neutrophils (Finisguerra et al. 2015). Therefore, MET amplification may have favored the recruitment of MPO-deficient neutrophils in the microenvironment of the mediastinal sarcoma. Indeed, the tumor presented a massive overexpression of pro-inflammatory cytokines, comprising TNFA, IFN-γ, IL6-Jak-STAT3, and IL2-STAT5 signaling cascade. Furthermore, our deconvolution of immune cell infiltrates clearly shows the enrichment of neutrophils, amongst other myeloid cells, in the microenvironment of the tumor. Therefore, the crosstalks between MET amplification and TNFA, as well as IFN-γ and IL-6 pathways (McLoughlin et al. 2003), may have fostered the recruitment of neutrophil in the tumor.

Chronic inflammation is a known feature of DS patients, driving interferonopathies and other autoinflammatory conditions (Huggard et al. 2020; Sullivan et al. 2017). In this patient, this baseline inflammatory condition may have been exacerbated by the predisposing splicing mutation on MPO. The inherited MPOD and the acquired genomic instability may have triggered proinflammatory pathways in the mediastinal sarcoma. Combined with the amplification of MET, the activation of proinflammatory signals have fostered the recruitment of MPO-impaired neutrophils, which likely could not have promoted cell death. Eventually, this condition may have had a role in the final chemoresistance and *exitus* of the patient.

## Methods

### Sample description

The tumor used in this study was collected from the patient before chemotherapy at the Regina Margherita Children’s Hospital (Turin). The patient was enrolled in the clinical trial entitled Genomic Profile Analysis in Children, Adolescents and Young Adult With Sarcomas - SAR_GEN-ITA (ClinicalTrials.gov ID: NCT04621201). The trial was approved on 30th November 2018 by the independent ethics committee of A.O.U. Città della Salute e della Scienza di Torino - A.O. Ordine Mauriziano - A.S.L. Città di Torino (Turin, Italy) and it was conducted according to the principles of the Declaration of Helsinki and Good clinical Practice. Parents were provided with written informed consent for the analysis and data publication.

### Fluorescence in situ hybridization

Validation of EWSR1 gene translocation (22q12.2) was performed through fluorescence in situ hybridization (FISH), using the ZytoLight SPEC EWSR1 Dual Color Break Apart Probe (ZytoVision GmbH, Bremerhaven, Germany) according to the manufacturer’s instructions. Red (ZyOrange, excitation 547 nm/emission 572 nm) and green (ZyGreen, excitation 503 nm/emission 528 nm) light probes targeted a proximal (chr 22:29,191,431-29,673,440) and a distal genomic (chr22:29,779,841-30,179,900) region near to the EWSR1 breakpoint. A 4 µm FFPE tumor slide was deparaffinized in xylene, de-masked using SCC (1x, pH 6) at 80°C for 20 min and digested with pepsin (0.5 mg ml−1 in 0.2 N HCl, pH 1.0; Protease and Protease Buffer II) (Abbott Laboratories, North Chicago, IL, US) for 17 min at 37 °C. Denaturation was then performed applying ten microlitres of probe onto each slide and placing them in a HYBrite (Abbott Laboratories) for 1 min at 85 °C, before overnight hybridization at 37°C. After multiple washings and counterstaining with DAPI, FISH signals were scored with an Olympus BX61 upright microscope, using a × 100 objective.

### Immunohistochemical assessment of tumor

A 3 µm slide was cut from a representative FFPE tumor block and immunohistochemistry was performed on a Ventana BenchMark ULTRA AutoStainer (Ventana Medical Systems, Tucson, AZ, USA) with the CD99 primary antibody (O13, mouse monoclonal antibody, prediluted, incubation time: 32 minutes, Ventana, Tucson, AZ, US). Antigen retrieval was performed using the CC1 antigen retrieval buffer (pH 8.5, EDTA, 100 °C, 52 min; Ventana Medical Systems, AZ, USA) and Ultraview was used to detect positivity through the chromogen 3, 3’ Diaminobenzidine (DAB). Nuclei were counterstained with Hematoxylin and Bluing reagent.

### DNA extraction and whole exome sequencing

Genomic DNA for the tumor was extracted from 10 μm-thick FFPE sections (3–6 sections per sample) using Maxwell® RSC DNA FFPE Kit (Promega Corporation) on Maxwell® RSC 48 Instrument (Promega Corporation) following the manufacturer’s protocol. Peripheral blood was used as a matching reference. DNA from blood samples was extracted with QIAamp DNA Blood Kit (QIAGEN) following the manufacturer’s protocol. Whole exome was captured from genomic DNA for tumor and matched normal using the SureSelect XT Human All Exon V6 + COSMIC (Agilent) following the manufacturer’s protocol as previously described (Cereda et al. 2016). Briefly, 0.2 μg of genomic DNA was subjected to hydrodynamic shearing by exposure to 3 minutes of sonication using a Covaris sonicator to obtain ∼200-bp-long fragments. Fragments were used to prepare libraries according to the SureSelect XT manual. Libraries were further amplified with 7–10 cycles of PCR and 150 ng were hybridized with the bait library. Captured DNA was amplified with 14 PCR cycles and barcode indexes were added. Libraries were sequenced using Illumina NovaSeq600 in 150nt-long paired-end modality.

### Sequence alignment and variant calling

Germline and somatic mutations were identified integrating our previously published pipeline (Cereda et al. 2016) with the GATK Best Practice guidelines as implemented in the HaTSPiL framework (Morandi et al. 2019). In particular, sequencing reads from each sample were aligned to the human genome reference (GRCh37/hg19) using Novoalign (http://www.novocraft.com/) with default parameters. At most three mismatches per read were allowed and PCR duplicates were removed using Picard Markduplicates tool (Broad Institute 2022). To improve accuracy of variant calling, local realignment around indels was performed using GATK RealignerTargetCreator and IndelRealigner tools. Single nucleotide variants (SBSs) and small insertion/deletions (IDs) were identified using MuTect v.1.1.17 (Cibulskis et al. 2013), Strelka v.1.0.15 (Saunders et al. 2012) and Varscan2 v.2.3.6 (Koboldt et al. 2012) in tumor and normal samples independently. Only variants identified as ‘KEEP’ and ‘PASS’ in MuTect and Strelka, respectively, were considered. SBSs and InDels were retained if (i) had allele frequency ≥5% and (ii) in a genomic position covered by at least 10 reads.

### Identification of inherited genomic aberration

Frequency distributions of the germline heterozygous single SNVs identified by varscan2 were inspected to assess chromosome aberrations in the inherited genome of the patient. As previously proposed (Cereda et al. 2016), in a diploid genome heterozygous SNVs follow a normal distribution centered around an allele frequency of 50% because both alleles are present at equal frequency in cells. In the case of allelic imbalance due to CNVs, the frequency distribution of heterozygous SNPs deviates from normality because of the unbalanced ratio between allele copies. Hence, the distribution of heterozygous SNP frequencies was used to confirm the presence of genomic alterations in the genome of the patient. To identify relevant germline mutations we selected SNPs that harbor an allele frequency ≥25%. Clinical interpretation of germline mutations was derived from ClinVar database (https://www.ncbi.nlm.nih.gov/clinvar/) and InterVar (Q. Li and Wang 2017), which exploits ACMG2015 guidelines (Richards et al. 2015), as previously described (Berrino et al. 2022). Mutations with Combined Annotation Dependent Depletion (CADD) (Kircher et al. 2014) PHREAD score higher or equal to 10 were considered as ‘deleterious’. Ensembl Variant Effect Predictor (McLaren et al. 2016) MaxEntScan (Yeo and Burge 2004; Shamsani et al. 2019) was used to predict pathogenic variant effects.

### Copy number detection and purity and ploidy estimation

Somatic CNV regions were identified using Sequenza v.3.0.0 (Favero et al. 2015) with parameters window=5mb and min.reads.baf=4, keeping only positions that are covered at least by 10 reads and EXCAVATOR2 (D’Aurizio et al. 2016) with binsize=20,000 and mode=paired. To identify amplified and deleted genes, the genomic coordinates of the aberrant regions were intersected with those of 20,297 human protein coding genes of the GENCODE GRCh37 version 28 (Frankish et al. 2019). A gene was considered as modified if ⩾80% of its length was contained in an aberrant region. Sequenza was also used to estimate purity and ploidy values.

### Identification of cancer driver mutations

In the tumor sample, SBSs and InDels from the three different tools were identified as somatic if absent in the normal counterpart. ANNOVAR (K. Wang, Li, and Hakonarson 2010) was used to identify nonsilent (i.e. nonsynonymous, stopgain, stoploss, frameshift, nonframeshift and splicing modifications) mutations using RefSeq v.64 (http://www.ncbi.nlm.nih.gov/RefSeq/) as a reference protein dataset. SBSs and InDels falling within 2 bp from the splice sites of a gene in one of the three datasets were considered as splicing mutations. Next, a list of cancer genes was retrieved from the Network of Cancer Genes v.5 (An et al. 2016) (http://ncg.kcl.ac.uk/). This list was exploited to select 183 and 518 pediatric and adult cancer driver genes, respectively. Of these, 23 and 63 were pediatric and adult sarcoma driver genes, respectively (Supplementary Table 8). Furthermore, a list of 164 genes with actionable alterations was collected from the ‘PrecisionTrialDrawer’ R package (Melloni et al. 2018) and considered as actionable genes (Supplementary Table 8). Genes harboring nonsilent mutations were annotated using these two gene lists. All nonsilent mutations but frameshift substitutions were retained if (i) identified by at least two variant callers or (ii) in genes annotated as cancer driver and/or actionable. Mutations with Combined Annotation Dependent Depletion (CADD) (Kircher et al. 2014) PHREAD score higher or equal to 20 were considered as ‘highly deleterious’. CancerVar (Q. Li et al. 2022) was used to classify the pathogenicity of somatic variants according to AMP/ASCO/CAP/CGC 2017-2019 guidelines (M. M. Li et al. 2017). Finally, variant frequencies were corrected by the tumor content reported by Sequenza.

### Mutational and CNV signature analysis

Mutational signature analyses were performed on all somatic mutations using SigProfilerMatrixGenerator (Bergstrom et al. 2019) and SigProfilerExtractor (Islam et al. 2022) as previously described (Thatikonda et al. 2023). Copy number signature analysis was performed on Sequenza results using R package ‘sigminer’ (S. Wang, Li, et al. 2021; S. Wang, Tao, et al. 2021) as previously described (S. Wang, Li, et al. 2021). Copy number burden was evaluated using the read_copynumber function from ‘sigminer’ (S. Wang, Li, et al. 2021; S. Wang, Tao, et al. 2021).

### Total RNA extraction and sequencing

Total RNA extracted from tumor biopsy using the RSC RNA FFPE Kit on Maxwell instrument. To exclude genomic contamination, total RNA was treated with DNAse I and cleared with RNA Clean and Concentration (Zymo Research). RNA quantity and quality were determined by Qubit Fluorometric Quantitation (Thermo Fisher Scientific) and using the RNA 6000 Nano kit on a Bioanalyzer (Agilent Technologies), respectively. RNA-seq library was generated from 0.1 µg of RNA using Illumina Total RNA Prep Stranded Ligation with Ribo-Zero according to manufacturer’s recommendations, and sequenced on Illumina NovaSeq6000 in 100nt-long paired-end read modality.

### Gene fusion and expression analyses of RNA-seq data

Raw sequencing reads were trimmed to avoid nucleotide overlaps between read pairs on both ends using the bbduck tool from bbmap (Bushnell 2014) v.38.18 with parameters forcetrimright=50 and minlength=30. Trimmed reads were aligned to the human genome reference GENCODE GRCh38 version 33 (Frankish et al. 2019) using STAR v.2.7.3a (Dobin et al. 2013) in basic two-pass mode removing duplicates and preventing multimappings (i.e. - -bamRemoveDuplicatesType UniqueIdentical and --outFilterMultimapNmax 1). Moreover, the following parameters were used: --alignInsertionFlush Right --outSAMstrandField intronMotif --outSAMattributes NH HI NM MD AS XS --peOverlapNbasesMin 20 --peOverlapMMp 0.25 -- chimSegmentMin 12 --chimJunctionOverhangMin 8 --chimOutJunctionFormat 1 -- chimMultimapScoreRange 3 --chimScoreJunctionNonGTAG -4 --chimMultimapNmax 20 and --chimNonchimScoreDropMin 10. Gene fusions were identified using STAR-Fusion v. 1.9.0 with options --min_FFPM 0 --FusionInspector validate --examine_coding_effect. Only fusions (FFPM≥0.1, LargeAnchorSupport=”YES”, LeftBreakEntropy≥1 and RightBreakEntropy≥1) were retained for further analysis. Read counts at gene level were estimated using featureCounts from Subread v. 2.0.0 (Liao, Smyth, and Shi 2014) with parameters -O –primary -Q 1 -J -s 2 -p -B. The number of transcripts per million reads (TPM) was measured starting from the expression values of 19,923 protein coding genes.

### Ontogeny signatures evaluation

Nine signatures related to ontogeny phases (namely endoderm, mesoderm, ectoderm, ectoderm early 1, ectoderm early 2, neural ectoderm anterior, neural ectoderm posterior, neuromesoderm progenitor early and neuromesoderm progenitor late) were retrieved from two publications (Messmer et al. 2019; Grosswendt et al. 2020). The mouse-derived ones (ectoderm early 1, ectoderm early 2, neural ectoderm anterior, neural ectoderm posterior, neuromesoderm progenitor early and neuromesoderm progenitor late) were converted to human gene symbols using the function gorth from the R package gprofiler2 v. 0.2.0 using as parameters source_organism=”mmusculus” and target_organism=”hsapiens”. Signatures were then grouped into 5 macrocategories according to their origin, namely Ectoderm (ectoderm, ectoderm early 1 and ectoderm early 2), Endoderm (endoderm), Mesoderm (mesoderm), Neuroectoderm (neural ectoderm anterior and neural ectoderm posterior) and Neuromesoderm (neuromesoderm progenitor early and neuromesoderm progenitor late). The expression in TPM of genes belonging to these categories was evaluated.

### Differential expression analysis

Gene expression data for EWS samples collected at diagnoses from three young (<4 years old) pediatric patients (i.e. SJEWS030998, SJEWS031029, SJEWS031208) available from the St.Jude Cloud (McLeod et al. 2021) were retrieved under acquired accession. Raw counts were normalized as transcript per million reads (TPM) using the human genome reference GENCODE GRCh38 version 33 (Frankish et al. 2019) as reference. Differential expression analysis was performed using the ‘edgeR’ R package(Robinson, McCarthy, and Smyth 2010) comparing the mediastinal EWS and the EWSs from the St.Jude database. Pvalues were corrected for multiple testing using Benjamini-Hochberg method (Benjamini and Hochberg 1995). Genes that presented an absolute log2(fold change)>1 and an adjusted pvalue≤0.1 were considered as differentially expressed.

### Over-representation analysis

Over representation analyses were performed with the enricher function in the R package ‘clusterProfileR’ (Yu et al. 2012; Wu et al. 2021) using either the 50 Hallmark, the 158 KEGG or the 278 positional gene sets defined in the mSigDb (Subramanian et al. 2005) and available through the R package ‘msigdbr’. Terms with pvalue≤0.05 were considered as significantly enriched. KEGG superfamilies of pathways were collected from the KEGG pathway databases (https://www.genome.jp/kegg/pathway.html).

### Definition of a list of neutrophil-related genes

A list of neutrophil-related genes was manually created on the basis of the work of Hendrick and Malanchi (Hedrick and Malanchi 2022) (Supplementary Table 8).

### Deconvolution of tumor tissue cellular heterogeneity

Normalized gene expression data (TPM) of the mediastinal EWS and the EWSs available form the St.Jude database were deconvolved using xCell into 64 cell-type-specific singature (Aran, Hu, and Butte 2017). In particular, xCellAnalysis function from the R package ‘xCell’ (https://github.com/dviraran/xCell) was used.

## Data Availability

All data produced in the present study are available upon reasonable request to the authors

## Acknowledgments

The research leading to these results has received funding from AIRC under MFAG 2017 ID 20566 (to M.C.), Ricerca Finalizzata 2019 ID GR-2019-12368827 (to M.C.), FPRC 5xmille 2018 Ministero Salute, project “ADVANCE/A-Bi-C”: Italian Ministry of Health, Ricerca Corrente 2021 (to M.C.), Compagnia di San Paolo (to M.C. and F.F.), and Fondazione Umberto Veronesi (to F.F.).

## Authors’ contribution

E.T., A.C., and M.C. conceived the study; E.T., S.P. and M.C. designed the analyses; K.M. and E.M. collected the samples; C.P. and S.G. performed sequencing experiments; S.P., F.P. and S.Per. run the bioinformatics analyses; S.P. and M.C. visualized the data; L.B. and M.P. performed pathological, immunohistochemical, and FISH analyses; E.T., S.D.A. and A.C. collected clinical information; E.T. and M.C. interpreted the data; M.C. supervised the study; M.C. and F.F. collected findings; E.T., S.P., F.F. and M.C. wrote the manuscript.

## References

1. Alexandrov, L. B., P. H. Jones, D. C. Wedge, J. E. Sale, P. J. Campbell, S. Nik-Zainal, and M. R. Stratton. 2015. “Clock-like Mutational Processes in Human Somatic Cells.” Nature Genetics 47 (12). 10.1038/ng.3441.

2. Alexandrov, L. B., J. Kim, N. J. Haradhvala, M. N. Huang, Tian Ng Aw, Y. Wu, A. Boot, et al. 2020. “The Repertoire of Mutational Signatures in Human Cancer.” Nature 578 (7793). 10.1038/s41586-020-1943-3.

3. An, O., G. M. Dall’Olio, T. P. Mourikis, and F. D. Ciccarelli. 2016. “NCG 5.0: Updates of a Manually Curated Repository of Cancer Genes and Associated Properties from Cancer Mutational Screenings.” Nucleic Acids Research 44 (D1). 10.1093/nar/gkv1123.

4. Aran, D., Z. Hu, and A. J. Butte. 2017. “xCell: Digitally Portraying the Tissue Cellular Heterogeneity Landscape.” Genome Biology 18 (1). 10.1186/s13059-017-1349-1.

5. Arbajian, E., J. Köster, Vult von Steyern F, and F. Mertens. 2018. “Inflammatory Leiomyosarcoma Is a Distinct Tumor Characterized by near-Haploidization, Few Somatic Mutations, and a Primitive Myogenic Gene Expression Signature.” Modern Pathology: An Official Journal of the United States and Canadian Academy of Pathology, Inc 31 (1). 10.1038/modpathol.2017.113.

6. Benjamini, Yoav, and Yosef Hochberg. 1995. “Controlling the False Discovery Rate: A Practical and Powerful Approach to Multiple Testing.” Journal of the Royal Statistical Society: Series B (Methodological). 10.1111/j.2517-6161.1995.tb02031.x.

7. Bergstrom, E. N., M. N. Huang, U. Mahto, M. Barnes, M. R. Stratton, S. G. Rozen, and L. B. Alexandrov. 2019. “SigProfilerMatrixGenerator: A Tool for Visualizing and Exploring Patterns of Small Mutational Events.” BMC Genomics 20 (1). 10.1186/s12864-019-6041-2.

8. Berrino, E., R. Filippi, C. Visintin, S. Peirone, E. Fenocchio, G. Farinea, F. Veglio, et al. 2022. “Collision of Germline POLE and PMS2 Variants in a Young Patient Treated with Immune Checkpoint Inhibitors.” NPJ Precision Oncology 6 (1). 10.1038/s41698-022-00258-8.

9. Bridge, J. A., J. R. Neff, D. A. Borek, and D. A. Hackbarth. 1990. “Primary Skeletal Ewing’s Sarcoma in Down Syndrome.” Cancer Genetics and Cytogenetics 47 (1). 10.1016/0165-4608(90)90263-a.

10. Broad Institute. 2022. “Picard Tools.” 2022. https://broadinstitute.github.io/picard/.

11. Bull, M. J. 2020. “Down Syndrome.” The New England Journal of Medicine 382 (24). 10.1056/NEJMra1706537.

12. Bushnell, Brian. 2014. “BBMap: A Fast, Accurate, Splice-Aware Aligner.” LBNL-7065E. Lawrence Berkeley National Lab. (LBNL), Berkeley, CA (United States). https://www.osti.gov/servlets/purl/1241166.

13. Casorzo, L., L. Fessia, A. Sapino, G. Ponzio, and G. Bussolati. 1989. “Extraskeletal Ewing’s Tumor with Translocation t(11;22) in a Patient with Down Syndrome.” Cancer Genetics and Cytogenetics 37 (1). 10.1016/0165-4608(89)90077-0.

14. Cereda, M., G. Gambardella, L. Benedetti, F. Iannelli, D. Patel, G. Basso, R. F. Guerra, et al. 2016. “Patients with Genetically Heterogeneous Synchronous Colorectal Cancer Carry Rare Damaging Germline Mutations in Immune-Related Genes.” Nature Communications 7 (July). 10.1038/ncomms12072.

15. Cibulskis, K., M. S. Lawrence, S. L. Carter, A. Sivachenko, D. Jaffe, C. Sougnez, S. Gabriel, M. Meyerson, E. S. Lander, and G. Getz. 2013. “Sensitive Detection of Somatic Point Mutations in Impure and Heterogeneous Cancer Samples.” Nature Biotechnology 31 (3). 10.1038/nbt.2514.

16. D’Aurizio, R., T. Pippucci, L. Tattini, B. Giusti, M. Pellegrini, and A. Magi. 2016. “Enhanced Copy Number Variants Detection from Whole-Exome Sequencing Data Using EXCAVATOR2.” Nucleic Acids Research 44 (20). 10.1093/nar/gkw695.

17. Davies, M. J., C. L. Hawkins, D. I. Pattison, and Rees. 2008. “Mammalian Heme Peroxidases: From Molecular Mechanisms to Health Implications.” Antioxidants & Redox Signaling 10 (7). 10.1089/ars.2007.1927.

18. Dobin, A., C. A. Davis, F. Schlesinger, J. Drenkow, C. Zaleski, S. Jha, P. Batut, M. Chaisson, and T. R. Gingeras. 2013. “STAR: Ultrafast Universal RNA-Seq Aligner.” Bioinformatics 29 (1). 10.1093/bioinformatics/bts635.

19. European Commission. 2018. “European Platform on Rare Disease Registration.” August 24, 2018. https://eu-rd-platform.jrc.ec.europa.eu.

20. Favero, F., T. Joshi, A. M. Marquard, N. J. Birkbak, M. Krzystanek, Q. Li, Z. Szallasi, and A. C. Eklund. 2015. “Sequenza: Allele-Specific Copy Number and Mutation Profiles from Tumor Sequencing Data.” Annals of Oncology: Official Journal of the European Society for Medical Oncology / ESMO 26 (1). 10.1093/annonc/mdu479.

21. Finisguerra, V., G. Di Conza, M. Di Matteo, J. Serneels, S. Costa, A. A. Thompson, E. Wauters, et al. 2015. “MET Is Required for the Recruitment of Anti-Tumoural Neutrophils.” Nature 522 (7556). 10.1038/nature14407.

22. Frankish, A., M. Diekhans, A. M. Ferreira, R. Johnson, I. Jungreis, J. Loveland, J. M. Mudge, et al. 2019. “GENCODE Reference Annotation for the Human and Mouse Genomes.” Nucleic Acids Research 47 (D1). 10.1093/nar/gky955.

23. Gröbner, S. N., B. C. Worst, J. Weischenfeldt, I. Buchhalter, K. Kleinheinz, V. A. Rudneva, P. D. Johann, et al. 2018. “The Landscape of Genomic Alterations across Childhood Cancers.” Nature 555 (7696). 10.1038/nature25480.

24. Grosswendt, S., H. Kretzmer, Z. D. Smith, A. S. Kumar, S. Hetzel, L. Wittler, S. Klages, B. Timmermann, S. Mukherji, and A. Meissner. 2020. “Epigenetic Regulator Function through Mouse Gastrulation.” Nature 584 (7819). 10.1038/s41586-020-2552-x.

25. Grünewald, T. G. P., F. Cidre-Aranaz, D. Surdez, E. M. Tomazou, E. de Álava, H. Kovar, P. H. Sorensen, O. Delattre, and U. Dirksen. 2018. “Ewing Sarcoma.” Nature Reviews. Disease Primers 4 (1). 10.1038/s41572-018-0003-x.

26. Hasle, H. 2001. “Pattern of Malignant Disorders in Individuals with Down’s Syndrome.” The Lancet Oncology 2 (7). 10.1016/S1470-2045(00)00435-6.

27. Hasle, H., J. M. Friedman, J. H. Olsen, and S. A. Rasmussen. 2016. “Low Risk of Solid Tumors in Persons with Down Syndrome.” Genetics in Medicine: Official Journal of the American College of Medical Genetics 18 (11). 10.1038/gim.2016.23.

28. Hedrick, C. C., and I. Malanchi. 2022. “Neutrophils in Cancer: Heterogeneous and Multifaceted.” Nature Reviews. Immunology 22 (3). 10.1038/s41577-021-00571-6.

29. Hicks, R. J., and E. W. Lau. 2009. “PET/MRI: A Different Spin from under the Rim.” European Journal of Nuclear Medicine and Molecular Imaging 36 Suppl 1 (March). 10.1007/s00259-008-0966-z.

30. Huggard, D., L. Kelly, E. Ryan, F. McGrane, N. Lagan, E. Roche, J. Balfe, et al. 2020. “Increased Systemic Inflammation in Children with Down Syndrome.” Cytokine 127 (March). 10.1016/j.cyto.2019.154938.

31. Islam, S. M. A., M. Díaz-Gay, Y. Wu, M. Barnes, R. Vangara, E. N. Bergstrom, Y. He, et al. 2022. “Uncovering Novel Mutational Signatures by de Novo Extraction with SigProfilerExtractor.” Cell Genomics 2 (11). 10.1016/j.xgen.2022.100179.

32. Kaul, T., C. Lotterman, and R. Warrier. 2019. “Adolescent With Down Syndrome Who Refuses to Walk.” Clinical Pediatrics 58 (11-12). 10.1177/0009922819868685.

33. Kircher, Martin, Daniela M. Witten, Preti Jain, Brian J. O’Roak, Gregory M. Cooper, and Jay Shendure. 2014. “A General Framework for Estimating the Relative Pathogenicity of Human Genetic Variants.” Nature Genetics 46 (3): 310.

34. Klebanoff, S. J. 2005. “Myeloperoxidase: Friend and Foe.” Journal of Leukocyte Biology 77 (5). 10.1189/jlb.1204697.

35. Koboldt, D. C., Q. Zhang, D. E. Larson, D. Shen, McLellan, L. Lin, C. A. Miller, E. R. Mardis, L. Ding, and R. K. Wilson. 2012. “VarScan 2: Somatic Mutation and Copy Number Alteration Discovery in Cancer by Exome Sequencing.” Genome Research 22 (3). 10.1101/gr.129684.111.

36. Kongkiatkamon, S., L. Terkawi, Y. Guan, V. Adema, M. Hasipek, T. Dombrovski, M. Co, et al. 2022. “Rare Germline Alterations of Myeloperoxidase Predispose to Myeloid Neoplasms.” Leukemia, June. 10.1038/s41375-022-01630-0.

37. Landrum, M. J., S. Chitipiralla, G. R. Brown, C. Chen, B. Gu, J. Hart, D. Hoffman, et al. 2020. “ClinVar: Improvements to Accessing Data.” Nucleic Acids Research 48 (D1). 10.1093/nar/gkz972.

38. Lee, P., R. Bhansali, S. Izraeli, N. Hijiya, and J. D. Crispino. 2016. “The Biology, Pathogenesis and Clinical Aspects of Acute Lymphoblastic Leukemia in Children with Down Syndrome.” Leukemia 30 (9). 10.1038/leu.2016.164.

39. Liao, Y., G. K. Smyth, and W. Shi. 2014. “featureCounts: An Efficient General Purpose Program for Assigning Sequence Reads to Genomic Features.” Bioinformatics 30 (7). 10.1093/bioinformatics/btt656.

40. Liberzon, A., C. Birger, H. Thorvaldsdóttir, M. Ghandi, J. P. Mesirov, and P. Tamayo. 2015. “The Molecular Signatures Database (MSigDB) Hallmark Gene Set Collection.” Cell Systems 1 (6). 10.1016/j.cels.2015.12.004.

41. Li, M. M., M. Datto, E. J. Duncavage, S. Kulkarni, N. I. Lindeman, S. Roy, A. M. Tsimberidou, et al. 2017. “Standards and Guidelines for the Interpretation and Reporting of Sequence Variants in Cancer: A Joint Consensus Recommendation of the Association for Molecular Pathology, American Society of Clinical Oncology, and College of American Pathologists.” The Journal of Molecular Diagnostics: JMD 19 (1). 10.1016/j.jmoldx.2016.10.002.

42. Lin, P. P., Y. Wang, and G. Lozano. 2011. “Mesenchymal Stem Cells and the Origin of Ewing’s Sarcoma.” Sarcoma 2011. 10.1155/2011/276463.

43. Li, Q., Z. Ren, K. Cao, M. M. Li, K. Wang, and Y. Zhou. 2022. “CancerVar: An Artificial Intelligence-Empowered Platform for Clinical Interpretation of Somatic Mutations in Cancer.” Science Advances 8 (18). 10.1126/sciadv.abj1624.

44. Li, Q., and K. Wang. 2017. “InterVar: Clinical Interpretation of Genetic Variants by the 2015 ACMG-AMP Guidelines.” American Journal of Human Genetics 100 (2). 10.1016/j.ajhg.2017.01.004.

45. Marchetti, C., P. Patriarca, G. P. Solero, F. E. Baralle, and M. Romano. 2004. “Genetic Characterization of Myeloperoxidase Deficiency in Italy.” Human Mutation 23 (5). 10.1002/humu.20027.

46. Ma, X., Y. Liu, Y. Liu, L. B. Alexandrov, M. N. Edmonson, C. Gawad, X. Zhou, et al. 2018. “Pan-Cancer Genome and Transcriptome Analyses of 1,699 Paediatric Leukaemias and Solid Tumours.” Nature 555 (7696). 10.1038/nature25795.

47. McLaren, W., L. Gil, S. E. Hunt, H. S. Riat, G. R. Ritchie, A. Thormann, P. Flicek, and F. Cunningham. 2016. “The Ensembl Variant Effect Predictor.” Genome Biology 17 (1). 10.1186/s13059-016-0974-4.

48. McLeod, C., A. M. Gout, X. Zhou, A. Thrasher, D. Rahbarinia, S. W. Brady, M. Macias, et al. 2021. “St. Jude Cloud: A Pediatric Cancer Genomic Data-Sharing Ecosystem.” Cancer Discovery 11 (5). 10.1158/2159-8290.CD-20-1230.

49. McLoughlin, R. M., J. Witowski, R. L. Robson, T. S. Wilkinson, S. M. Hurst, A. S. Williams, J. D. Williams, S. Rose-John, S. A. Jones, and N. Topley. 2003. “Interplay between IFN-Gamma and IL-6 Signaling Governs Neutrophil Trafficking and Apoptosis during Acute Inflammation.” The Journal of Clinical Investigation 112 (4). 10.1172/JCI17129.

50. Melloni, Giorgio E. M., Alessandro Guida, Giuseppe Curigliano, Edoardo Botteri, Angela Esposito, Maude Kamal, Christoph Le Tourneau, et al. 2018. “Precision Trial Drawer, a Computational Tool to Assist Planning of Genomics-Driven Trials in Oncology.” JCO Precision Oncology, August. 10.1200/PO.18.00015.

51. Messmer, T., F. von Meyenn, A. Savino, F. Santos, H. Mohammed, A. T. L. Lun, J. C. Marioni, and W. Reik. 2019. “Transcriptional Heterogeneity in Naive and Primed Human Pluripotent Stem Cells at Single-Cell Resolution.” Cell Reports 26 (4). 10.1016/j.celrep.2018.12.099.

52. Miller, R. W. 1969. “Childhood Cancer and Congenital Defects. A Study of U.S. Death Certificates during the Period 1960-1966.” Pediatric Research 3 (5). 10.1203/00006450-196909000-00001.

53. Morandi, E., M. Cereda, D. Incarnato, C. Parlato, G. Basile, F. Anselmi, A. Lauria, et al. 2019. “HaTSPiL: A Modular Pipeline for High-Throughput Sequencing Data Analysis.” PloS One 14 (10). 10.1371/journal.pone.0222512.

54. Mp, Mac Manus, R. J. Hicks, J. P. Matthews, A. McKenzie, D. Rischin, E. K. Salminen, and D. L. Ball. 2003. “Positron Emission Tomography Is Superior to Computed Tomography Scanning for Response-Assessment after Radical Radiotherapy or Chemoradiotherapy in Patients with Non-Small-Cell Lung Cancer.” Journal of Clinical Oncology: Official Journal of the American Society of Clinical Oncology 21 (7). 10.1200/JCO.2003.07.054.

55. Nižetić, D., and J. Groet. 2012. “Tumorigenesis in Down’s Syndrome: Big Lessons from a Small Chromosome.” Nature Reviews. Cancer 12 (10). 10.1038/nrc3355.

56. Osuna-Marco, M. P., M. López-Barahona, B. López-Ibor, and Á. M. Tejera. 2021. “Ten Reasons Why People With Down Syndrome Are Protected From the Development of Most Solid Tumors -A Review.” Frontiers in Genetics 12 (November). 10.3389/fgene.2021.749480.

57. Pedersen R, S., G. Karemore, T. Gudjonsson, M. B. Rask, B. Neumann, J. K. Hériché, R. Pepperkok, et al. 2016. “Profiling DNA Damage Response Following Mitotic Perturbations.” Nature Communications 7 (December). 10.1038/ncomms13887.

58. Richards, S., N. Aziz, S. Bale, D. Bick, S. Das, J. Gastier-Foster, W. W. Grody, et al. 2015. “Standards and Guidelines for the Interpretation of Sequence Variants: A Joint Consensus Recommendation of the American College of Medical Genetics and Genomics and the Association for Molecular Pathology.” Genetics in Medicine: Official Journal of the American College of Medical Genetics 17 (5). 10.1038/gim.2015.30.

59. Robinson, D. J. McCarthy, and G. K. Smyth. 2010. “edgeR: A Bioconductor Package for Differential Expression Analysis of Digital Gene Expression Data.” Bioinformatics 26 (1). 10.1093/bioinformatics/btp616.

60. Satgé, D., A. J. Sasco, N. L. Carlsen, C. A. Stiller, H. Rubie, B. Hero, B. de Bernardi, et al. 1998. “A Lack of Neuroblastoma in Down Syndrome: A Study from 11 European Countries.” Cancer Research 58 (3). https://pubmed.ncbi.nlm.nih.gov/9458088/.

61. Satgé, D., A. J. Sasco, A. Chompret, D. Orbach, F. Méchinaud, B. Lacour, B. Roullet, et al. 2003. “A 22-Year French Experience with Solid Tumors in Children with Down Syndrome.” Pediatric Hematology and Oncology 20 (7). 10.1080/08880010390232727.

62. Satgé, D., C. A. Stiller, S. Rutkowski, A. O. von Bueren, B. Lacour, D. Sommelet, M. Nishi, et al. 2013. “A Very Rare Cancer in Down Syndrome: Medulloblastoma. Epidemiological Data from 13 Countries.” Journal of Neuro-Oncology 112 (1). 10.1007/s11060-012-1041-y.

63. Saunders, C. T., W. S. Wong, S. Swamy, J. Becq, L. J. Murray, and R. K. Cheetham. 2012. “Strelka: Accurate Somatic Small-Variant Calling from Sequenced Tumor-Normal Sample Pairs.” Bioinformatics 28 (14). 10.1093/bioinformatics/bts271.

64. Schwartz, L. H., S. Litière, E. de Vries, R. Ford, S. Gwyther, S. Mandrekar, L. Shankar, et al. 2016. “RECIST 1.1-Update and Clarification: From the RECIST Committee.” European Journal of Cancer 62 (July). 10.1016/j.ejca.2016.03.081.

65. Shamsani, J., S. H. Kazakoff, I. M. Armean, W. McLaren, M. T. Parsons, B. A. Thompson, T.A. O’Mara, S. E. Hunt, N. Waddell, and A. B. Spurdle. 2019. “A Plugin for the Ensembl Variant Effect Predictor That Uses MaxEntScan to Predict Variant Spliceogenicity.” Bioinformatics 35 (13). 10.1093/bioinformatics/bty960.

66. Strauss, S. J., A. M. Frezza, N. Abecassis, J. Bajpai, S. Bauer, R. Biagini, S. Bielack, et al. 2021. “Bone Sarcomas: ESMO-EURACAN-GENTURIS-ERN PaedCan Clinical Practice Guideline for Diagnosis, Treatment and Follow-Up.” Annals of Oncology: Official Journal of the European Society for Medical Oncology / ESMO 32 (12). 10.1016/j.annonc.2021.08.1995.

67. Subramanian, A., P. Tamayo, V. K. Mootha, S. Mukherjee, B. L. Ebert, M. A. Gillette, A. Paulovich, et al. 2005. “Gene Set Enrichment Analysis: A Knowledge-Based Approach for Interpreting Genome-Wide Expression Profiles.” Proceedings of the National Academy of Sciences of the United States of America 102 (43). 10.1073/pnas.0506580102.

68. Sullivan, K. D., D. Evans, A. Pandey, T. H. Hraha, K. P. Smith, N. Markham, A. L. Rachubinski, et al. 2017. “Trisomy 21 Causes Changes in the Circulating Proteome Indicative of Chronic Autoinflammation.” Scientific Reports 7 (1). 10.1038/s41598-017-13858-3.

69. Suster, D. I. 2020. “The Role of Molecular Pathology in Mediastinal Sarcomas.” Mediastinum (Hong Kong, China) 4 (December). 10.21037/med-20-39.

70. Thatikonda, V., S. M. A. Islam, R. J. Autry, B. C. Jones, S. N. Gröbner, G. Warsow, B. Hutter, et al. 2023. “Comprehensive Analysis of Mutational Signatures Reveals Distinct Patterns and Molecular Processes across 27 Pediatric Cancers.” Nature Cancer, January. 10.1038/s43018-022-00509-4.

71. Tirtei, E., M. Cereda, E. De Luna, P. Quarello, S. D. Asaftei, and F. Fagioli. 2020. “Omic Approaches to Pediatric Bone Sarcomas.” Pediatric Blood & Cancer 67 (2). 10.1002/pbc.28072.

72. Walther, C., M. Mayrhofer, J. Nilsson, J. Hofvander, T. Jonson, N. Mandahl, I. Øra, D. Gisselsson, and F. Mertens. 2016. “Genetic Heterogeneity in Rhabdomyosarcoma Revealed by SNP Array Analysis.” Genes, Chromosomes & Cancer 55 (1). 10.1002/gcc.22285.

73. Wang, G., W. Huang, S. Wang, J. Wang, W. Cui, W. Zhang, A. Lou, S. Geng, and X. Li. 2021. “Macrophagic Extracellular Vesicle CXCL2 Recruits and Activates the Neutrophil CXCR2/PKC/NOX4 Axis in Sepsis.” Journal of Immunology 207 (8). 10.4049/jimmunol.2100229.

74. Wang, K., M. Li, and H. Hakonarson. 2010. “ANNOVAR: Functional Annotation of Genetic Variants from High-Throughput Sequencing Data.” Nucleic Acids Research 38 (16). 10.1093/nar/gkq603.

75. Wang, S., H. Li, M. Song, Z. Tao, T. Wu, Z. He, X. Zhao, K. Wu, and X. S. Liu. 2021. “Copy Number Signature Analysis Tool and Its Application in Prostate Cancer Reveals Distinct Mutational Processes and Clinical Outcomes.” PLoS Genetics 17 (5). 10.1371/journal.pgen.1009557.

76. Wang, S., Z. Tao, T. Wu, and X. S. Liu. 2021. “Sigflow: An Automated and Comprehensive Pipeline for Cancer Genome Mutational Signature Analysis.” Bioinformatics 37 (11). 10.1093/bioinformatics/btaa895.

77. Wood, G. E., H. Hockings, D. M. Hilton, and S. Kermorgant. 2021. “The Role of MET in Chemotherapy Resistance.” Oncogene 40 (11). 10.1038/s41388-020-01577-5.

78. Wu, T., E. Hu, S. Xu, M. Chen, P. Guo, Z. Dai, T. Feng, et al. 2021. “clusterProfiler 4.0: A Universal Enrichment Tool for Interpreting Omics Data.” Innovation (Cambridge (Mass.)) 2 (3). 10.1016/j.xinn.2021.100141.

79. Yeo, G., and C. B. Burge. 2004. “Maximum Entropy Modeling of Short Sequence Motifs with Applications to RNA Splicing Signals.” Journal of Computational Biology: A Journal of Computational Molecular Cell Biology 11 (2-3). 10.1089/1066527041410418.

80. Yu, G., L. G. Wang, Y. Han, and Q. Y. He. 2012. “clusterProfiler: An R Package for Comparing Biological Themes among Gene Clusters.” Omics: A Journal of Integrative Biology 16 (5). 10.1089/omi.2011.0118.

81. Zöllner, S. K., J. F. Amatruda, S. Bauer, S. Collaud, E. de Álava, S. G. DuBois, J. Hardes, et al. 2021. “Ewing Sarcoma-Diagnosis, Treatment, Clinical Challenges and Future Perspectives.” Journal of Clinical Medicine Research 10 (8). 10.3390/jcm10081685.

