## Supplementary Figures for "Impaired neutrophil-mediated cell death drives Ewing’s Sarcoma in a two years old child with Down Syndrome" (from another version of this preprint: version 1, 2023-05-31)

### Supplementary Information

**Supplementary Figure 1. Genomic alterations characterizing the mediastinal EWS.**

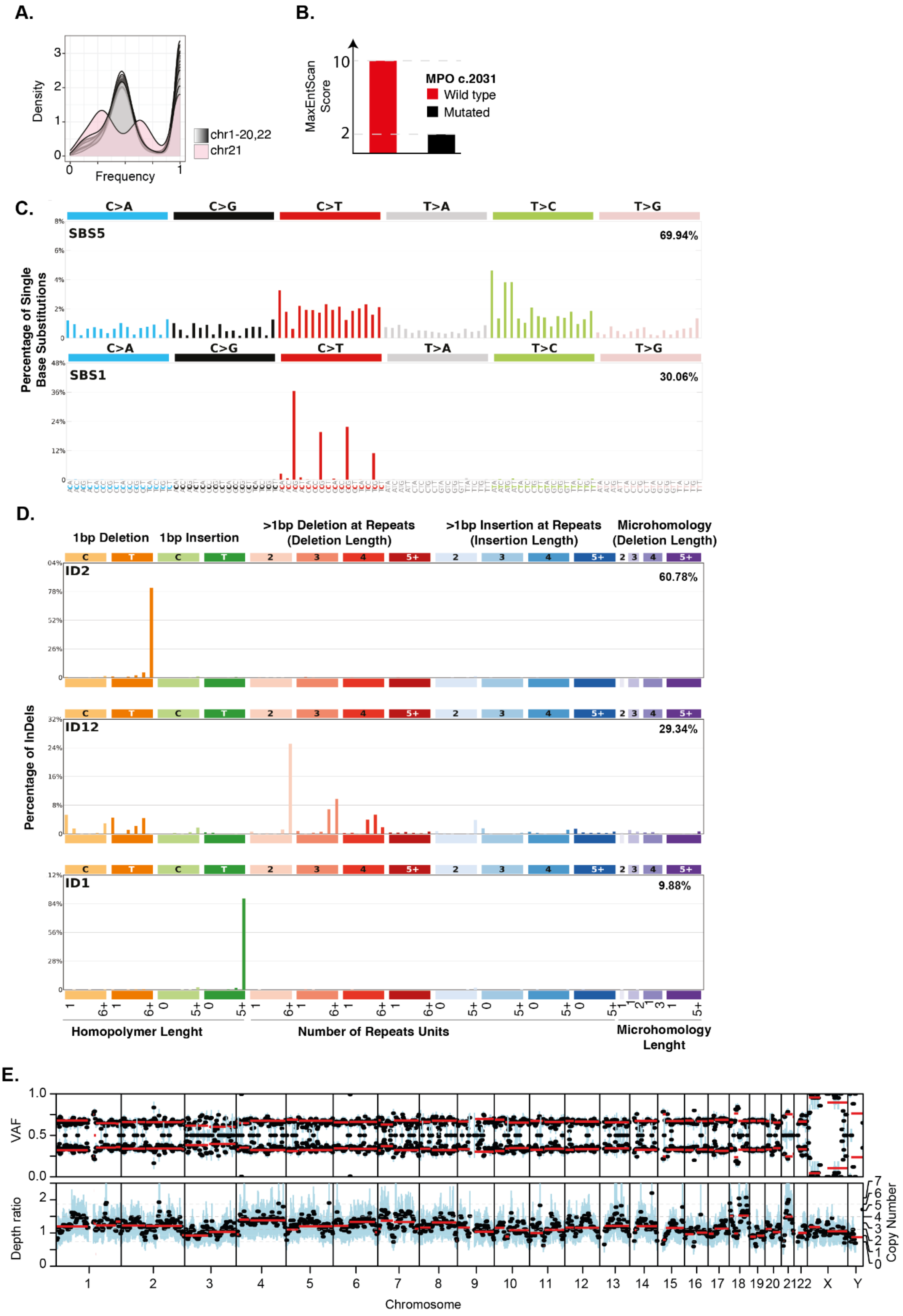

(A) Density distribution of variant allele frequency (VAF) of germline variants stratified by chromosomes. Chromosomes 1-20,22 are depicted in gray, whereas chromosome 21 is depicted in pink. (B) Bar plot shows MaxEntScan score measured for 3' splice sites at intron 11 and exon 12 junction encompassing the MPO c.2031-2A>C mutations. (C-D) Results of COSMIC SBS (C) and ID (D) signature deconvolution analyses. (E) Genome view represents VAF and the depth ratio profile of germline variants. Red horizontal lines represent mean value considering genome segments optimized by Sequenza.

#### Supplementary Figure 2. Transcriptomic analyses of pediatric EWSs.

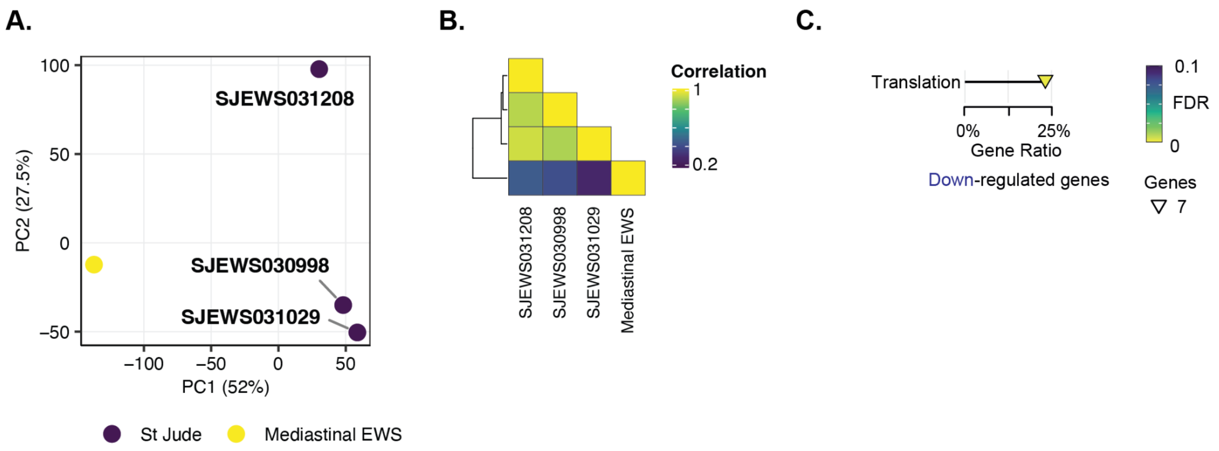


(A) Principal component analysis represents transcriptional distribution of the mediastinal EWS" and three EWS samples from young euploid children available at the St Jude portal (namely "St Jude"). (B) Heatmap shows pairwise correlations of gene expression levels across samples. (C) Over representation analysis performed on downregulated genes using KEGG gene sets.
